# Differential impact of antiretroviral therapy initiated before or during pregnancy on placenta pathology in HIV-positive women

**DOI:** 10.1101/2020.06.15.20131615

**Authors:** Nadia M Ikumi, Thokozile R Malaba, Komala Pillay, Marta C Cohen, Hlengiwe P Madlala, Mushi Matjila, Dilly Anumba, Landon Myer, Marie-Louise Newell, Clive M Gray, for the PIMS Study Group

**Author notes:** **Corresponding Author** Clive M Gray, Division of Immunology, Institute of Infectious Disease and Molecular Medicine, Falmouth Building, Faculty of Health Sciences, University of Cape Town, Anzio Road, Observatory, Cape Town, South Africa, 7925. Co-First Authors (Nadia M Ikumi, Thokozile R Malaba). **Conflict of interest statement** The authors report no conflict of interest.

## Abstract

**Objective:** To examine the association between timing of ART initiation in HIV-infected women and placental histopathology.

**Design:** A nested sub-study in a larger cohort of HIV-infected women which examined the association between ART status and birth outcomes.

**Methods:** Placentas (n=130) were examined for histopathology from two ART groups: *stable* (n=53), who initiated ART before conception and *initiating* (n=77), who started ART during pregnancy (median [IQR] 15 weeks gestation [11-18]). Using binomial regression we quantified associations between ART initiation timing with placental histopathology and pregnancy outcomes.

**Results:** One-third of all placentas were <10^th^ percentile weight-for-gestation and there was no significant difference between ART groups. Placental diameter, thickness, cord insertion position and fetal-placental weight ratio were also similar by group. However, placentas from the stable group showed increased maternal vascular malperfusion (MVM) (39.6% vs 19.4%), and decreased weight (392g vs 422g, p=0.09). MVM risk was twice as high (RR 2.03 [95% CI: 1.16 - 3.57]; p=0.01) in the stable group; the increased risk remaining significant when adjusting for maternal age (RR 2.04 [95% CI: 1.12 - 3.72]; p=0.02). Furthermore, MVM was significantly associated with pre-term delivery (PTD) and low birth weight (LBW) (p=0.002 and p<0.0001, respectively).

**Conclusion:** Preconception initiation of ART was associated with an increased MVM risk, and may contribute to placental dysfunction. The association between MVM with PTD and LBW suggests that a placenta-mediated mechanism likely links the putative association between long-term use of ART and adverse birth outcomes.

## Introduction

Antiretroviral treatment (ART) during pregnancy is not only of benefit for both the mother and infant, substantially decreasing MTCT, but is also proven to be most effective in reducing maternal and child mortality.^[1,2]^ Like many other countries, South Africa has now fully adopted universal ART for life for all HIV-positive women upon diagnosis, including pregnant women.^[3]^ However, the extensive rollout and uptake of ART from before or early pregnancy has led to a rapid rise in fetal exposure to ART; where such *in-utero* drug exposure has been associated with increased risk of adverse birth outcomes: including preterm delivery (PTD)^[4–6]^, low birth weight (LBW)^[7]^ and small-for-gestational age (SGA) infants^[4,8]^.

HIV infection in pregnancy, even in the absence of ART, has been associated with an increased risk of adverse birth outcomes which is thought to be through inflammation-mediated disruption of placental development and function^[9,10]^. How maternal HIV infection impacts placental structure and function is thus important to consider as it can affect both maternal health, as seen in eclampsia^[11]^, and fetal health, as seen with PTD and SGA^[9,12–14]^. Uncharacteristic placental shapes and sizes may indicate decreased placental efficiency, and maternal supply-line deprivation^[15,16]^. Histologic chorioamnionitis reflects maternal and fetal inflammatory responses to ascending infection^[17,18]^, and is a major cause of spontaneous PTD^[19–21]^. Despite recognition of the importance of the placenta in adverse birth outcomes, there is scant data on the impact of *in-utero* ART exposure on placental development and pathology, and subsequent adverse birth outcomes.

Within a large cohort of HIV-positive women, we set out to systematically examine the histopathology of placentas collected at delivery from HIV-positive women initiating ART before or during pregnancy, to inform understanding of the impact of ART initiation timing on placental pathology. We conjectured that initiation of ART during pregnancy would result in greater inflammation-linked placental pathology than ART initiated before pregnancy.

## Methods

We performed a nested sub-study in the Prematurity Immunology in HIV-infected Mothers and their infants Study (PIMS) cohort in Cape Town, South Africa^[22]^. PIMS was a prospective cohort study investigating the association between ART use and adverse birth outcomes, in particular PTD, SGA and LBW^[22]^. HIV-positive pregnant women aged ≥18 years seeking antenatal care (ANC) services at the Gugulethu Midwife Obstetric Unit (MOU) were recruited between April 2015 and October 2016, at ≤24 weeks gestation. Whole placentas were collected at delivery and only those graded as good quality (grades 1-3)^[23]^ were processed within 24 hours. Included were women who initiated ART before pregnancy (stable group), or during pregnancy (initiating group). ART regimens consisted predominately of tenofovir (TDF) + lamivudine (3TC)/emtricitabine (FTC) + efavirenz (EFV)^[22]^.

### Ethical considerations

The PIMS study protocol, informed consent forms and all data collection tools were approved by the University of Cape Town’s, Faculty of Health Sciences Human Research Ethics Committee (UCT FHS-HREC) (HREC 739/2014) and the University of Southampton Faculty of Medicine Ethics Committee on (12542 PIMS). Written informed consent was obtained from all study participants, including placenta collection for the nested cohort.

### Data collection

At first ANC visit, gestational age (GA) was determined by ultrasound scan (USS) by a research sonographer using a standardized assessment protocol. Obstetric outcomes, including date of delivery and birthweight, were obtained from clinical records. Maternal CD4 and viral load measurements were obtained from hospital records at the time closest to the first ANC visit. Body Mass Index (BMI) was calculated from weight and height measurements; blood pressure were measured at the first ANC visit (baseline) and during follow-up.

### Placenta collection and processing

Whole placentas were collected at delivery and fixed in 10% buffered formalin. Placental histopathological examination were performed in the Department of Pathology, Red Cross Memorial Children’s Hospital, Cape Town, South Africa. Specimens were examined from the umbilical cord, membranes and placental disk and scored following the Amsterdam Placental Workshop Group Consensus Statement^[24]^. Briefly, four blocks were prepared from each placenta, one block included a roll of the extraplacental membranes and two cross sections of the umbilical cord, one from the placental insertion, and one close to the fetal end. In addition, three blocks each contained a full-thickness section of the placenta parenchyma, one from a visibly abnormal area if present and two sections from macroscopically normal tissue including one from the cord insertion. The tissue sections were 3-5μm thick and were cut and stained with hematoxylin and eosin for histological examination. Macroscopic and microscopic features were combined to report placenta pathology. For the sake of pathology scoring rigour, sections were also assesed in the Department of Histopathology, Sheffield Children Hospital NHS Foundation Trust, Sheffield, UK.

### Variables of interest

Placenta pathology was based on placenta weight-for-gestational age, fetal-placental weight ratio and features of placenta pathology. All placenta weights were reported as trimmed weights, after removal of umbilical cords and membranes. Placenta weight percentiles by gestational age and fetal-placenta ratios were categorized as small (<10^th^), appropriate (10-90^th^) and large (>90^th^) for gestational age.^[25]^ Microscopically, the placenta was examined for features of meconium exposure, based on the presence and amount of meconium-laden macrophages in the placental membranes, with some cases showing features of prolonged exposure characterized by meconium deep in the chorionic plate and umbilical cord; amnion degeneration and chorion vascular muscle necrosis^[26]^. Histologic chorioamnionitis was defined as the presence of acute inflammation in the placenta, with or without a fetal inflammatory response^[27,28]^. Maternal vascular malperfusion (MVM) was defined as a constellation of macroscopic and histologic features representing a pattern of injury in the placenta as a result of maternal vascular abnormalities. The presence of decidual vasculopathy was indicative of maternal vascular malperfusion. In the absence of decidual vasculopathy, MVM was indicated by a small placenta for gestational age accompanied by at least two villous changes (villous infarct, retroplacental haematoma, accelerated villous maturation or distal villous hypoplasia). In the case of normal placenta weights for gestational age, at least 3-4 villous features were noted to be placed in the category of MVM.^[24,29,30]^ Decidual arteriopathy was defined as abnormal or incomplete remodelling of decidual spiral arterioles resulting in fibrinoid necrotic lesions^[24,29,30]^. Chronic deciduitis was defined as abnormal infiltration of lymphocytes and plasma cells in the decidua^[24,31]^. Placenta disk dimensions were recorded as maximum linear dimension (*length*), greatest dimension of the axis perpendicular to the *length* (*width*) and the thickness. Cord insertion referred to site of insertion on the placenta in relation to the margin of the placenta classified as central, off-centre and marginal based on the distance from the margin.

Secondary outcomes included PTD, SGA and LBW. PTD was defined as delivery at <37 weeks’ GA^[32]^. Birthweight was categorized as low (LBW, <2500g), normal (NBW, 2500-4000g) and high (HBW, >4000g)^[33]^. Size for gestational age, based on gender-specific INTERGROWTH-21^st^ project standards, was defined as small for gestational age (SGA) for infants with birthweight <10^th^ percentile for gestational age, appropriate for gestational age (AGA) for those between 10-90^th^ percentile and large for gestational age (LGA) for those >90^th^ percentiles^[34]^.

### Statistical analysis

Statistical analyses were performed using STATA version 12.0 (Stata Corporation, College Station, Texas, USA) and R. Analyses focused on placenta pathology outcomes by ART initiation before or during pregnancy. The impact of ART timing on features of placental pathology was summarized using an Alluvial plot generated using the *ggalluvial* package in R^[35]^. Secondary analyses focused on birth outcomes (PTD, LBW and SGA) related to features of placenta pathology. In bivariable analyses, proportions were compared using chi-squared/Fishers exact test and rank-sum tests. Associations between baseline characteristics and placenta pathology outcomes (small placental weight for GA, MVM and chorioamnionitis) were assessed using binomial regression. Results were presented as risk ratios (RR) with 95% confidence intervals (CI) generated using the forestplot package in R (R package version 1.9)^[36]^. For secondary analyses of birth outcomes, given the limited sample size, only bivariate associations were assessed. As placenta weight and MVM abnormalities were likely to originate in early gestation, we explored associations between timing of ART initiation and the placenta pathologies. For MVM, as advanced maternal age has been shown to increase the risk of placental dysfunction,^[37]^ we applied a multivariable regression model to adjust for maternal age differences recorded at booking between ART groups. To assess potential risk factors for PTD, SGA and LBW we compared differences between groups using the chi^2^ test or Fisher’s exact test. Missing values were excluded from these analyses.

## Results

A total of 130 placentas were examined: 53 (40.8%) from the stable group and 77 (59.2%) from the initiating group of women. Maternal and obstetric characteristics are summarised in Table 1.

**Table 1:**
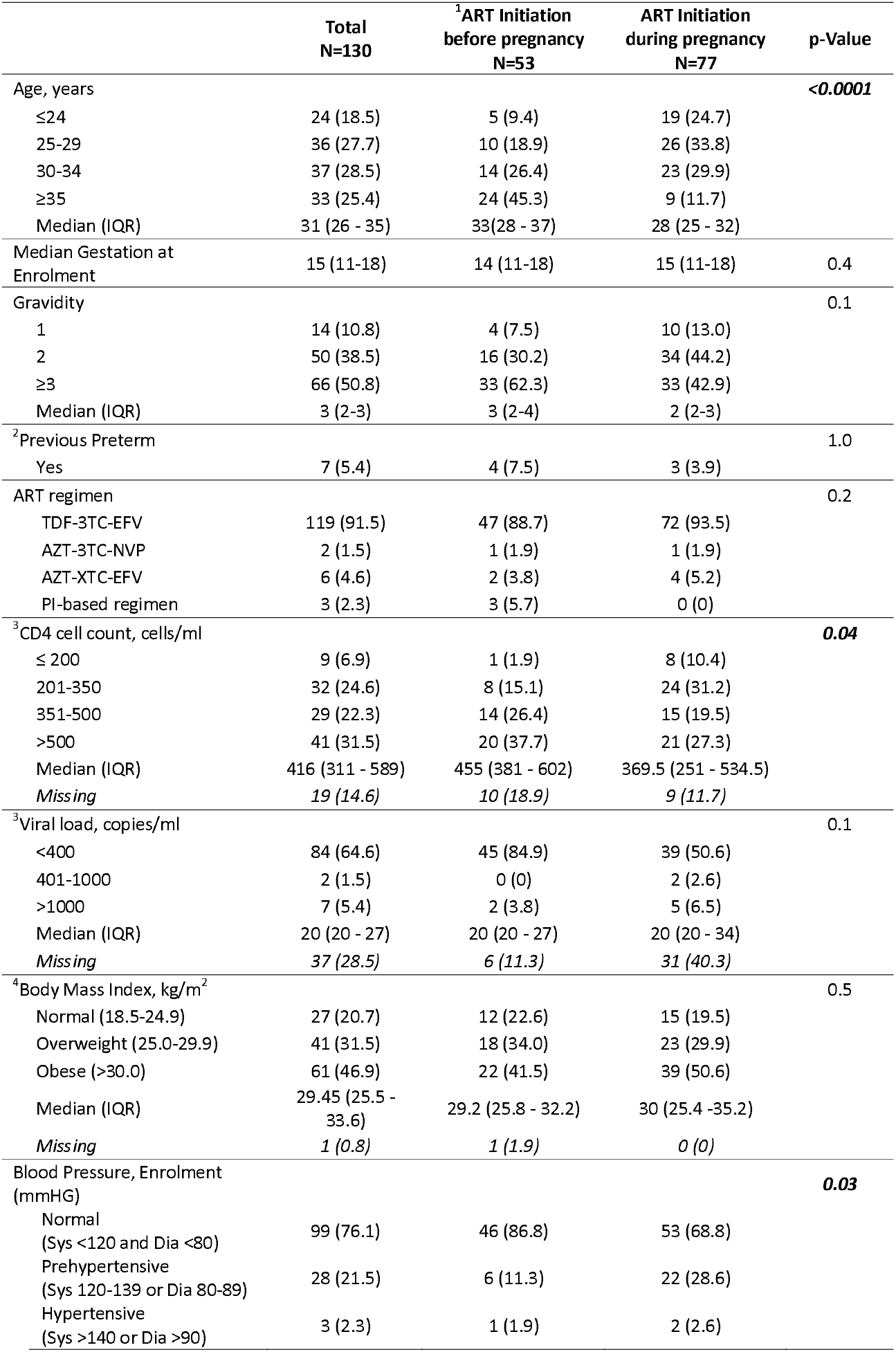
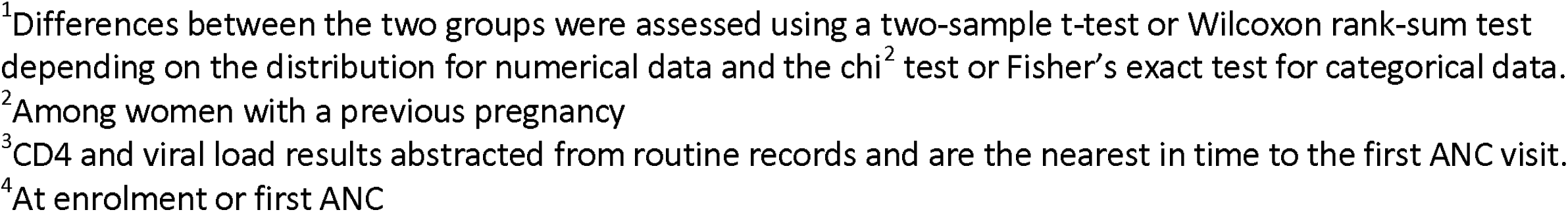
Maternal and obstetric characteristics of the pregnant women at enrolment.

|  | Total<br>N=130 | <sup>1</sup>ART Initiation<br>before pregnancy<br>N=53 | ART Initiation<br>during pregnancy<br>N=77 | p-Value |
| --- | --- | --- | --- | --- |
| Age, years |  |  |  | **<0.0001** |
| ≤24 | 24 (18.5) | 5 (9.4) | 19 (24.7) |  |
| 25-29 | 36 (27.7) | 10 (18.9) | 26 (33.8) |  |
| 30-34 | 37 (28.5) | 14 (26.4) | 23 (29.9) |  |
| ≥35 | 33 (25.4) | 24 (45.3) | 9 (11.7) |  |
| Median (IQR) | 31 (26 - 35) | 33 (28 - 37) | 28 (25 - 32) |  |
| Median Gestation at<br>Enrolment | 15 (11-18) | 14 (11-18) | 15 (11-18) | 0.4 |
| Gravidity |  |  |  | 0.1 |
| 1 | 14 (10.8) | 4 (7.5) | 10 (13.0) |  |
| 2 | 50 (38.5) | 16 (30.2) | 34 (44.2) |  |
| ≥3 | 66 (50.8) | 33 (62.3) | 33 (42.9) |  |
| Median (IQR) | 3 (2-3) | 3 (2-4) | 2 (2-3) |  |
| <sup>2</sup>Previous Preterm |  |  |  | 1.0 |
| Yes | 7 (5.4) | 4 (7.5) | 3 (3.9) |  |
| ART regimen |  |  |  | 0.2 |
| TDF-3TC-EFV | 119 (91.5) | 47 (88.7) | 72 (93.5) |  |
| AZT-3TC-NVP | 2 (1.5) | 1 (1.9) | 1 (1.9) |  |
| AZT-XTC-EFV | 6 (4.6) | 2 (3.8) | 4 (5.2) |  |
| PI-based regimen | 3 (2.3) | 3 (5.7) | 0 (0) |  |
| <sup>3</sup>CD4 cell count, cells/ml |  |  |  | **0.04** |
| ≤ 200 | 9 (6.9) | 1 (1.9) | 8 (10.4) |  |
| 201-350 | 32 (24.6) | 8 (15.1) | 24 (31.2) |  |
| 351-500 | 29 (22.3) | 14 (26.4) | 15 (19.5) |  |
| >500 | 41 (31.5) | 20 (37.7) | 21 (27.3) |  |
| Median (IQR) | 416 (311 - 589) | 455 (381 - 602) | 369.5 (251 - 534.5) |  |
| Missing | 19 (14.6) | 10 (18.9) | 9 (11.7) |  |
| <sup>3</sup>Viral load, copies/ml |  |  |  | 0.1 |
| <400 | 84 (64.6) | 45 (84.9) | 39 (50.6) |  |
| 401-1000 | 2 (1.5) | 0 (0) | 2 (2.6) |  |
| >1000 | 7 (5.4) | 2 (3.8) | 5 (6.5) |  |
| Median (IQR) | 20 (20 - 27) | 20 (20 - 27) | 20 (20 - 34) |  |
| Missing | 37 (28.5) | 6 (11.3) | 31 (40.3) |  |
| <sup>4</sup>Body Mass Index, kg/m<sup>2</sup> |  |  |  | 0.5 |
| Normal (18.5-24.9) | 27 (20.7) | 12 (22.6) | 15 (19.5) |  |
| Overweight (25.0-29.9) | 41 (31.5) | 18 (34.0) | 23 (29.9) |  |
| Obese (>30.0) | 61 (46.9) | 22 (41.5) | 39 (50.6) |  |
| Median (IQR) | 29.45 (25.5 - 33.6) | 29.2 (25.8 - 32.2) | 30 (25.4 - 35.2) |  |
| Missing | 1 (0.8) | 1 (1.9) | 0 (0) |  |
| Blood Pressure, Enrolment<br>(mmHG) |  |  |  | **0.03** |
| Normal<br>(Sys <120 and Dia <80) | 99 (76.1) | 46 (86.8) | 53 (68.8) |  |
| Prehypertensive<br>(Sys 120-139 or Dia 80-89) | 28 (21.5) | 6 (11.3) | 22 (28.6) |  |
| Hypertensive<br>(Sys >140 or Dia >90) | 3 (2.3) | 1 (1.9) | 2 (2.6) |  |

<sup>1</sup>Differences between the two groups were assessed using a two-sample t-test or Wilcoxon rank-sum test depending on the distribution for numerical data and the $\chi^2$ test or Fisher's exact test for categorical data. <sup>2</sup>Among women with a previous pregnancy <sup>3</sup>CD4 and viral load results abstracted from routine records and are the nearest in time to the first ANC visit. <sup>4</sup>At enrolment or first ANC

### Maternal characteristics

Women in the stable group were older (median 33 years, IQR 28-37) than initiating women (median 28, IQR 25-32). Median GA at first ANC visit was 15 weeks (IQR 11-18), with no differences by ART initiation timing group (Table 1). Overall, 119 (91.5%), women were on a fixed dose regimen of Tenofovir (TDF) + Lamivudine (3TC) + Efavirenz (EFV), 2 (1.5%) were on Zidovudine (AZT) + Lamividune (3TC) + Nevirapine (NVP), 6 (4.6%) were on AZT + Emtricitabine or Lamivudine (XTC) + EFV with no difference between the groups; only 3 (2.3%; all in the *stable* group) were on a protease inhibitor (PI)-based regimen. Stable women had significantly higher median CD4 T cell counts at enrolment (455 cells/ml, IQR 381 - 602) than initiating women (369.5 cells/ml, IQR 251 - 534.5). Most women (78.4%) were overweight or obese, not significantly different by ART group. Initiating women were more likely to be pre-hypertensive than stable women (28.6% vs 11.3%). Alcohol was the most used substance, with 22% of women reporting alcohol use < 30 days prior to first ANC visit (Supp. Table 1). There was no difference in smoking or recreational drug use between the two groups.

### Infant characteristics

Of the 130 live singleton births, 58 (44.6%) were female and 72 (55.3%) male (Table 2), with no sex difference by ART timing group. Overall, median gestation at delivery was 39 weeks (IQR 38 - 40), and PTD incidence was low (6.9%) with no difference by timing of ART initiation. While median birthweight was also similar between ART groups, there was a tendency towards differences in the percentage of LBW (11.3% vs 6.5%) and HBW (1.9% vs 9.1%) deliveries in the stable and initiating group respectively; but not reaching statistical significance. The tendency towards increased >4000g macrosomia births in the ART initiation during pregnancy group was not associated with birth complications. Differences were also observed in size for gestational age, where 12 (9.2%) infants were SGA overall, with 8 (15.1%) in the stable and 4 (5.2%) in the initiating group.

**Table 2:** Infant characteristics

|  | Total<br>N=130 | ART Initiation before<br>pregnancy<br>N=53 | ART Initiation<br>during pregnancy<br>N=77 | p-Value |
| --- | --- | --- | --- | --- |
| Sex |  |  |  | 0.6 |
| Female | 58 (44.6) | 25 (47.2) | 33 (42.9) |  |
| Male | 72 (55.4) | 28 (52.8) | 44 (57.1) |  |
| Median Delivery Gestation<br>(completed weeks) | 39 (38 - 40) | 39 (38 - 39) | 39 (38 - 40) | 0.1 |
| Birthweight, grams |  |  |  | 0.2 |
| Low birthweight (<2500g) | 11 (8.5) | 6 (11.3) | 5 (6.5) |  |
| Normal (2500 - 4000g) | 111 (85.4) | 46 (86.8) | 65 (84.4) |  |
| High (>4000g) | 8 (6.1) | 1 (1.9) | 7 (9.1) |  |
| Median (IQR) | 3180 (2880 - 3540) | 3140 (2690 - 3360) | 3240 (2940 - 3580) |  |
| Preterm (<37 weeks<br>Gestational Age) |  |  |  | 1.0 |
| Yes | 9 (6.9) | 4 (7.5) | 5 (6.5) |  |
| No | 121 (93.1) | 49 (92.5) | 72 (93.5) |  |
| Size for Gestational Age, <10<sup>th</sup><br>centile |  |  |  | 0.07 |
| Yes | 12 (9.2) | 8 (15.1) | 4 (5.2) |  |
| No | 118 (90.8) | 45 (84.9) | 73 (94.8) |  |
| Mode of Delivery |  |  |  | 0.2 |
| Vaginal Delivery | 97 (74.6) | 42 (79.2) | 55 (71.4) |  |
| Elective Caesarean | 24 (18.5) | 4 (7.6) | 4 (5.2) |  |
| Emergency Caesarean | 8 (6.1) | 6 (11.3) | 18 (23.4) |  |
| Missing | 1 (0.8) | 1 (1.9) | 0 (0) |  |

<sup>1</sup>Differences between the two groups were assessed using a two-sample t-test or Wilcoxon rank-sum test depending on the distribution for numerical data and the chi<sup>2</sup> test or Fisher's exact test for categorical data.

**Table 4:** Relationship between placenta pathology and preterm delivery, size for gestational age and low birthweight.

|  | <sup>1</sup>Preterm delivery |  |  | <sup>1</sup>Size for gestational age (<10<sup>th</sup> centile) |  |  | <sup>1</sup>Low birthweight (<2500g) |  |  |
| --- | --- | --- | --- | --- | --- | --- | --- | --- | --- |
|  | Y (%) | N (%) | p | Y (%) | N (%) | p | Y (%) | N (%) | p |
| **Percentile placental weight** |  |  | 0.7 |  |  | 0.1 |  |  | 1.0 |
| <10<sup>th</sup> | 2 (23) | 41 (34) |  | 7 (58) | 36 (30) |  | 4 (36) | 39 (33) |  |
| >10<sup>th</sup> | 6 (67) | 80 (66) |  | 5 (42) | 81 (69) |  | 7 (64) | 79 (66) |  |
| Missing | 1 (11) | 0 (0) |  | 0(0) | 1(1) |  | 0 (0) | 1(1) |  |
| **Cord insertion** |  |  | 0.2 |  |  | 1.0 |  |  | 0.1 |
| Central | 3 (33) | 17 (14) |  | 2 (17) | 18 (15) |  | 4 (36) | 16 (13) |  |
| Off centre | 5 (56) | 83 (69) |  | 8 (67) | 80 (68) |  | 7 (64) | 81 (68) |  |
| Marginal | 0 (0) | 16 (13) |  | 1 (8) | 15 (13) |  | 0 (0) | 16 (14) |  |
| Missing | 1 (11) | 5 (4) |  | 1 (8) | 5 (4) |  | 0 (0) | 6 (5) |  |
| **Meconium exposure** |  |  | 1.0 |  |  | 0.01 |  |  | 0.7 |
| + | 1 (11) | 24 (19) |  | 6 (50) | 19 (16) |  | 1 (9) | 24 (20) |  |
| - | 8 (89) | 97 (80) |  | 6 (50) | 99 (84) |  | 10 (91) | 95 (78) |  |
| **Chorioamnionitis** |  |  |  |  |  |  |  |  |  |
| *Maternal Inflammatory Response* |  |  | 0.4 |  |  | 0.4 |  |  | 0.4 |
| + | 0 (0) | 19 (16) |  | 3 (25) | 16 (14) |  | 0 | 19 (16) |  |
| - | 9 (100) | 102 (84) |  | 9 (75) | 102 (86) |  | 11 (100) | 100 (84) |  |
| *Fetal Inflammatory Response* |  |  | 0.4 |  |  | 1.0 |  |  | 0.2 |
| + | 0 | 20 (17) |  | 2 (17) | 18 (15) |  | 0 | 20 (17) |  |
| - | 9 (100) | 101 (83) |  | 10 (83) | 100 (85) |  | 11 (100) | 99 (83) |  |
| **Chronic Deciduitis** |  |  | 1.0 |  |  | 0.4 |  |  | 0.4 |
| + | 1 (11) | 13 (11) |  | 0 | 14 (12) |  | 2 (18) | 12 (10) |  |
| - | 8 (89) | 108 (89) |  | 12 (100) | 104 (88) |  | 9 (82) | 107 (90) |  |
| **Decidual arteriopathy** |  |  | 0.07 |  |  | 0.5 |  |  | 0.1 |
| + | 2 (22) | 5 (4) |  | 1 (8) | 6 (5) |  | 2 (18) | 5 (4) |  |
| - | 7 (78) | 116 (96) |  | 11 (92) | 112 (95) |  | 9 (82) | 114 (96) |  |
| **Maternal Vascular Malperfusion (MVM)** |  |  | 0.002 |  |  | 0.09 |  |  | <0.0001 |
| + | 7 (78) | 29 (24) |  | 6 (50) | 30 (25) |  | 10 (91) | 26 (22) |  |
| - | 2 (23) | 92 (76) |  | 6 (50) | 88 (75) |  | 1 (9) | 93 (78) |  |

<sup>1</sup>To assess potential risk factors for preterm delivery (<37 weeks gestational age), size for gestational age (<10<sup>th</sup> percentile) and low birth weight (<2500g), we compared differences between groups using the chi<sup>2</sup> test or Fisher's exact test.

### Placental Pathology

Table 3 shows placenta characteristics and pathology stratified by timing of ART initiation, and Figures 1A-D show representative images of some of the features of placenta pathology included in our analysis: chorioamnionitis (Figure 1A), neutrophilic infiltrate involving the cord vein representing a fetal response (Figure 1B) and decidual artery atherosis (Figure 1C). Median placenta weight was 412g (IQR 350 - 480) overall, 393g in the stable and 422g in the initiating group (p=0.09). Overall, 43 (33%) placentas were <10^th^ percentile weight-for-gestation, with no significant differences between ART groups; placental diameter, thickness or cord insertion were also similar in each group as was the Fetal-Placental weight ratio. Meconium exposure was identified in 22 (19.2%) cases, similar in both ART groups. Chorioamnionitis was present in 19 (14.6%) cases, with a higher percentage in initiating than in stable women, but not reaching statistical significance. MVM was significantly higher in placentas from stable than initiating women (39.2% vs 19.5%; p=0.01). There were no cases of chronic basal villitis, chronic chorioamnionitis, eosinophilic/T-cell vasculitis or chronic histiocytic intervillositis. Chronic decidual perivasculitis was included in cases of chronic deciduitis.

**Table 3:** Placental characteristics and pathology at delivery among women initiating ART before or during pregnancy.

|  | Total<br>N=130 | <sup>1</sup>ART Initiation<br>before pregnancy<br>N=53 | ART Initiation<br>during pregnancy<br>N=77 | P-value |
| --- | --- | --- | --- | --- |
| Placental weight (g), median (IQR) | 412 (346 - 480) | 392 (346 – 440) | 422 (344 – 490) | 0.09 |
| Placental weight (g) |  |  |  | 1.0 |
| Small (<10<sup>th</sup> percentile) | 43 (33.1) | 17 (32.1) | 26 (33.8) |  |
| Appropriate (10 – 90<sup>th</sup> percentile) | 85 (65.4) | 35 (66.0) | 50 (64.9) |  |
| Large (>90<sup>th</sup> percentile) | 1 (0.8) | 0 (0) | 1 (1.3) |  |
| Missing | 1 (0.8) | 1 (1.9) | 0 (0) |  |
| Fetal-Placenta weight ratio |  |  |  | 0.9 |
| Small (<10<sup>th</sup> percentile) | 3 (2.3) | 1 (1.9) | 2 (2.6) |  |
| Appropriate (10 – 90<sup>th</sup> percentile) | 81 (62.3) | 32 (60.4) | 49 (63.6) |  |
| Large (>90<sup>th</sup> percentile) | 45 (34.6) | 19 (35.8) | 26 (33.8) |  |
| Missing | 1 (0.8) | 1 (1.9) | 0 (0) |  |
| Placenta greatest diameter (mm), median (IQR) | 170 (160 – 180) | 175 (161 – 189) | 170 (160 – 180) | 0.1 |
| Placenta thickness (mm), median (IQR) | 17.5 (10 – 25) | 15 (10 – 22) | 20 (10 - 25) | 0.4 |
| Cord insertion |  |  |  | 0.1 |
| Central | 20 (15.4) | 6 (11.3) | 14 (18.2) |  |
| Off centre | 88 (67.7) | 34 (64.1) | 54 (70.1) |  |
| Marginal | 16 (12.3) | 10 (18.9) | 6 (7.8) |  |
| Missing | 6 (4.6) | 3 (5.7) | 3 (3.9) |  |
| **Meconium exposure** |  |  |  | 0.9 |
| + | 25 (19.2) | 10 (18.9) | 15 (19.5) |  |
| - | 105 (80.8) | 43 (81.1) | 62 (80.5) |  |
| **Chorioamnionitis** |  |  |  |  |
| *Maternal Inflammatory Response* |  |  |  | 0.8 |
| + | 19 (14.6) | 7 (13.2) | 12 (15.6) |  |
| - | 111 (85.4) | 46 (86.3) | 65 (84.4) |  |
| *Fetal Inflammatory Response* |  |  |  | 0.1 |
| + | 20 (15.4) | 5 (9.4) | 15 (19.5) |  |
| - | 110 (84.6) | 48 (90.6) | 62 (80.5) |  |
| **Chronic Deciduitis** |  |  |  | 0.8 |
| + | 14 (10.8) | 5 (9.4) | 9 (11.7) |  |
| - | 116 (89.2) | 48 (90.6) | 68 (88.3) |  |
| **Maternal Vascular Malperfusion (MVM)** |  |  |  | **0.01** |
| + | 36 (27.7) | 21 (39.6) | 15 (19.5) |  |
| - | 94 (72.3) | 32 (60.4) | 62 (80.5) |  |

<sup>1</sup>Differences between the two groups were assessed using a two-sample t-test or Wilcoxon rank-sum test depending on the distribution for numerical data and the chi<sup>2</sup> test or Fisher's exact test for categorical data.

**Figure 1A:**
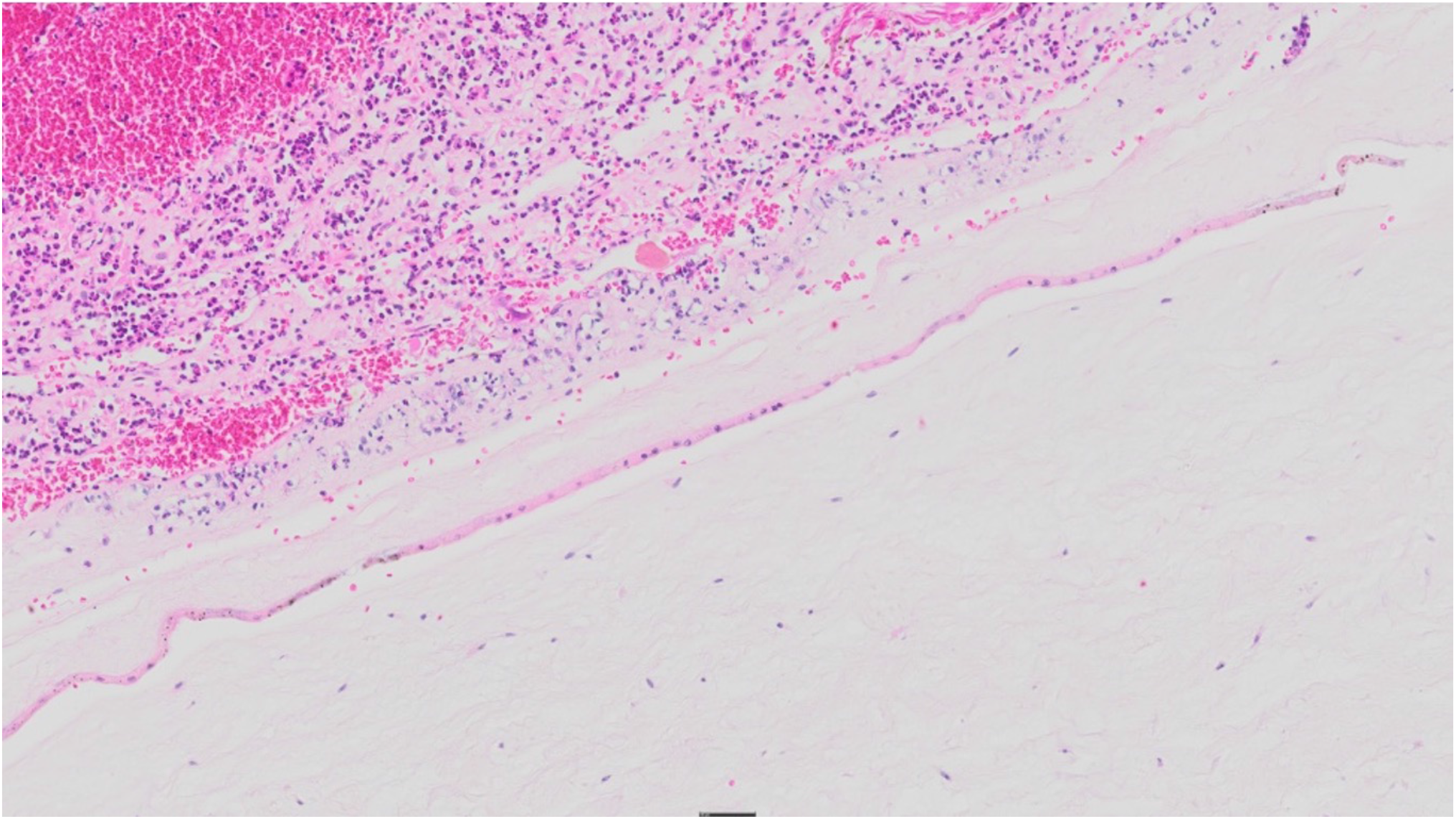
Digital scan of a representative Haematoxylin and Eosin (H&E) stained section showing chorioamnionitis of the membranes (maternal response).

**Figure 1B:**
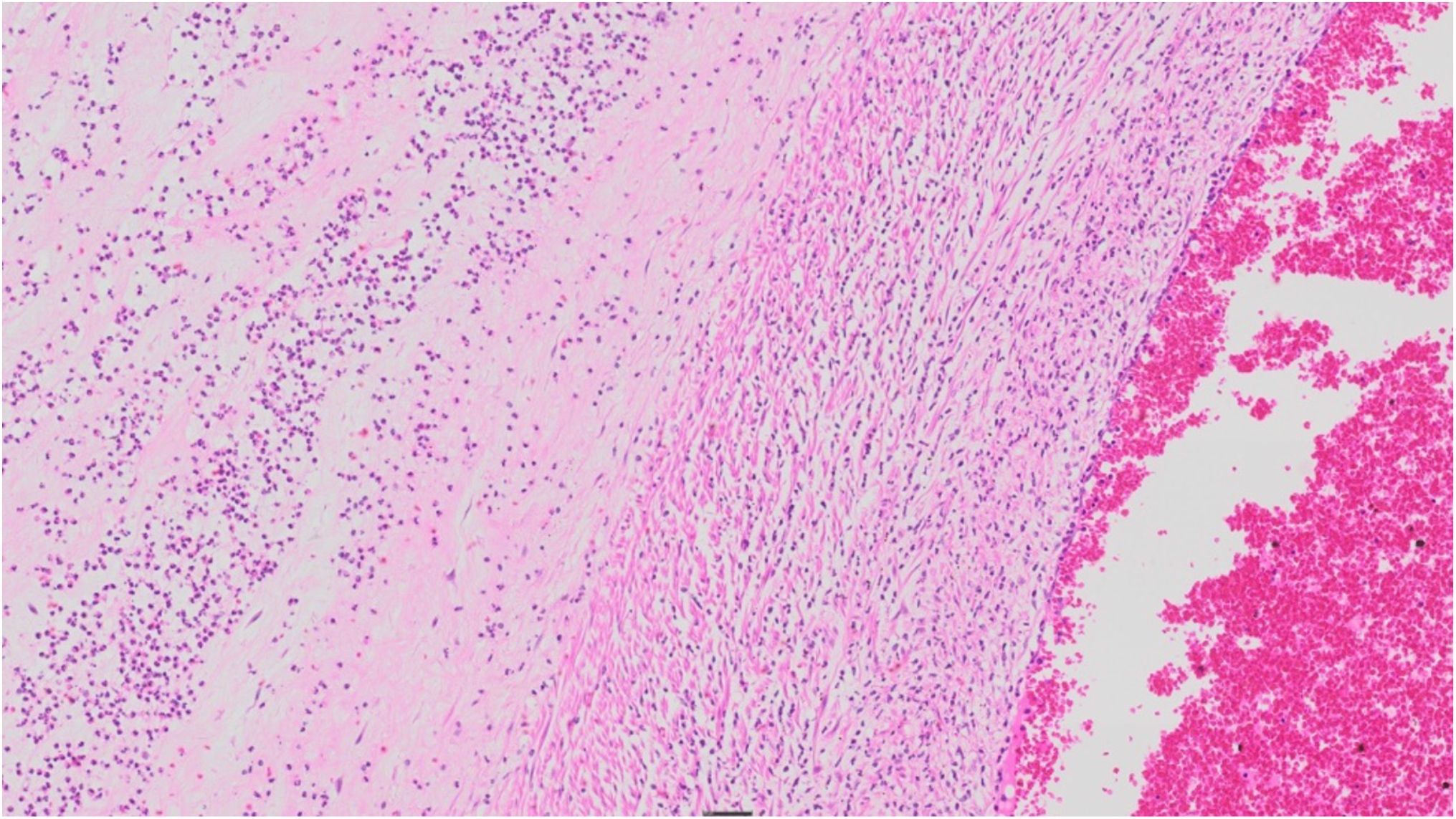
Digital scan of a representative H&E stained section showing a neutrophilic infiltrate involving the cord vein wall extending into Wharton’s jelly of the cord (fetal response).

**Figure 1C:**
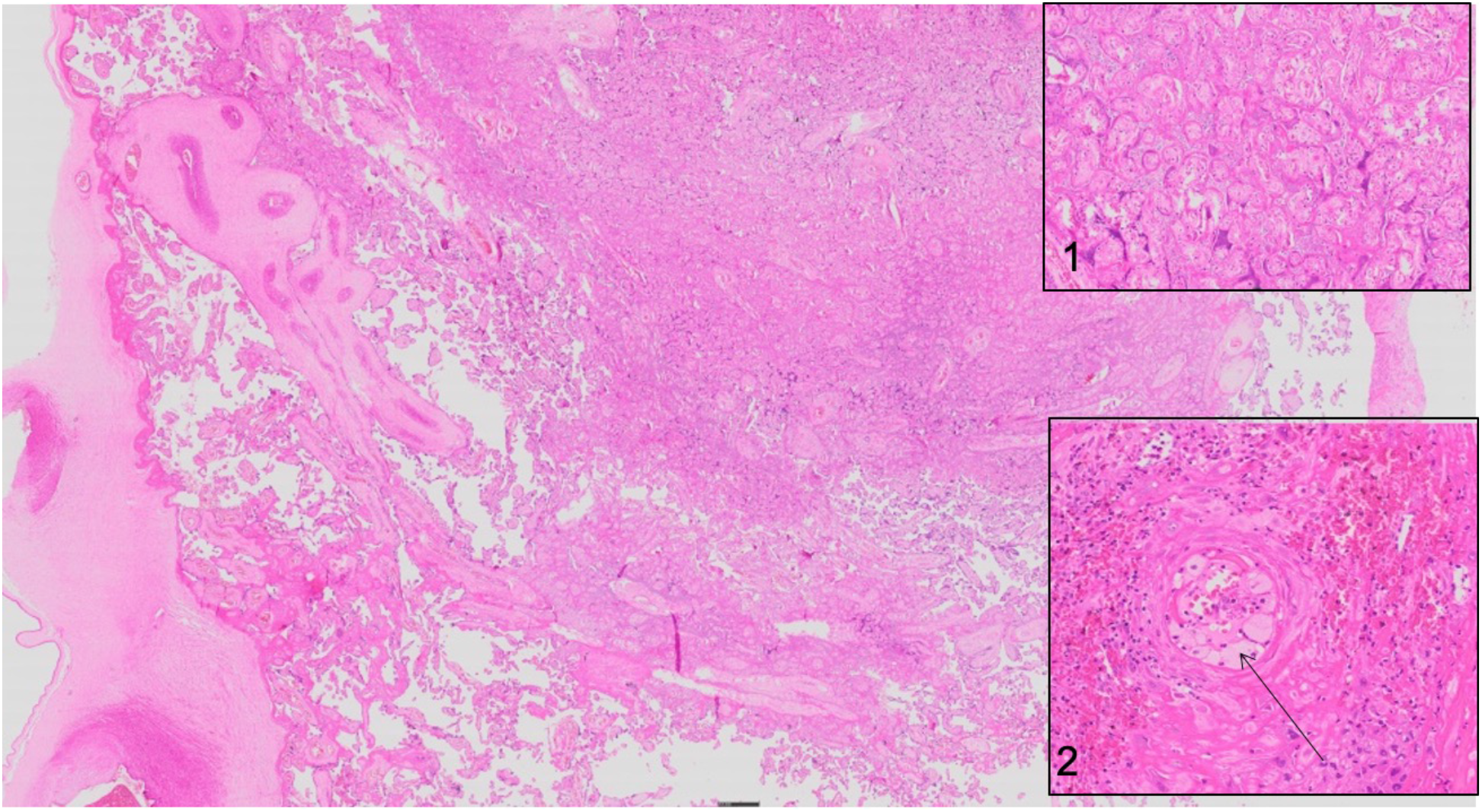
Digital scans of representative H&E stained sections of the placenta showing an infarct. Insert 1 shows a magnified image of the infarct and Insert 2 shows a magnified foam cell (denoted by the arrow) within the decidual artery demonstrating atherosis.

To discount selection bias due to collection of placentas from a sub-group of women, we compared maternal characteristics in “no-placenta” versus “placenta” collection for each ART group (Supp Table 2) in the larger PIMS cohort. Importantly, blood pressure was no different in the wider cohort (n=421), irrespective of ART group, indicating that the observed MVM in the smaller placenta stable group was not due to any bias in a pre- or hypertensive state, which could have impacted on our observations.

Figures 2A-C shows the relative risk of placenta pathology by ART group and maternal characteristics. In unadjusted models, women who were overweight (RR 0.46 [95% CI: 0.27-081]; p=0.007) and obese (RR 0.37 [95% CI: 0.21 - 0.64]; p<0.0001) were less likely to have small-for-gestational age placentas (Figure 2A). None of the variables explored were associated with significant risk of chorioamionitis (Figure 2B). The risk of MVM (Figure 2C) was twice as high (RR 2.03 [95% CI: 1.16 - 3.57]; p=0.01) in stable than in initiating women and remained significant when adjusting for maternal age (RR 2.04 [95% CI: 1.12 - 3.72]; p=0.02). The impact of ART timing on placental pathology is summarized in the Alluvial plot in Figure 2D, emphasizing the occurrence of MVM in women initiating ART pre-conception. The association between placenta pathology and PTD, SGA and LBW (Table 4) was significant for MVM (PTD: p=0.002; LBW: p<0.0001), and borderline for SGA (p=0.09). In addition, SGA deliveries were more likely to present with meconium exposure, indicating fetal distress (p=0.01).

**Figure 2A:**
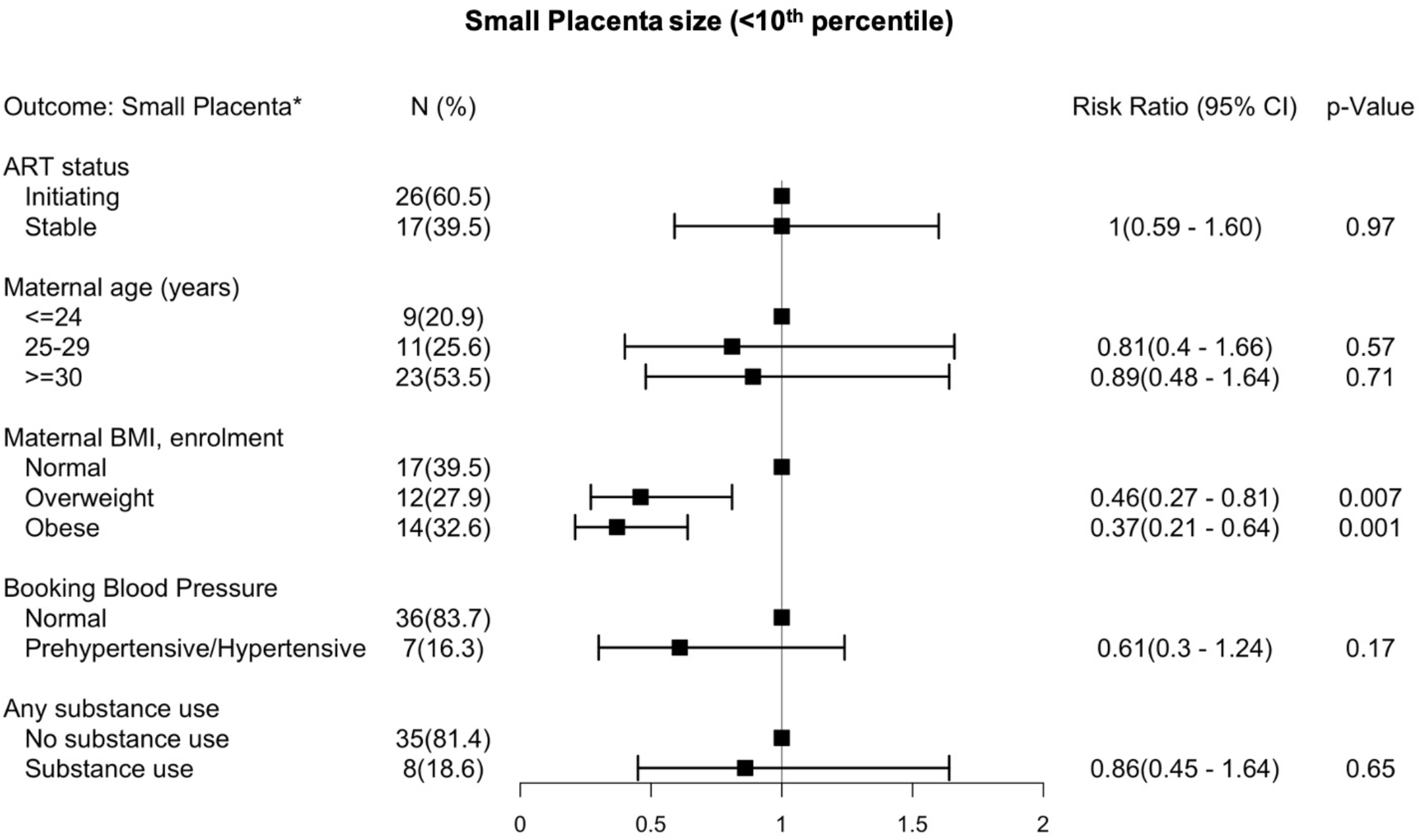
Forest plot depicting the relative risk with the 95% confidence intervals of HIV infected women presenting with a small placenta (<10^th^ percentile for gestational age).

**Figure 2B:**
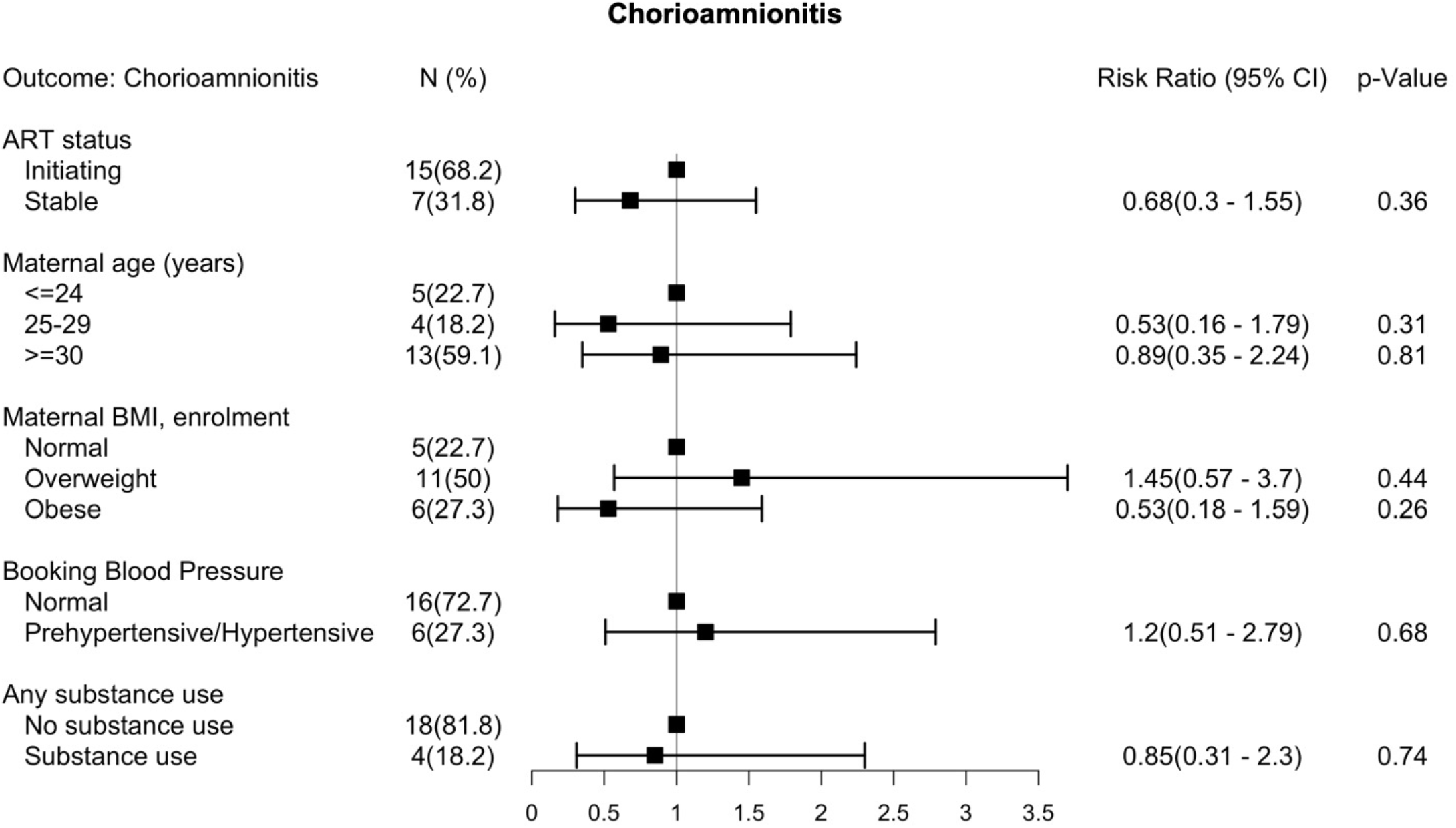
Forest plot depicting the relative risk with the 95% confidence intervals of HIV infected women presenting histologic chorioamnionitis.

**Figure 2C:**
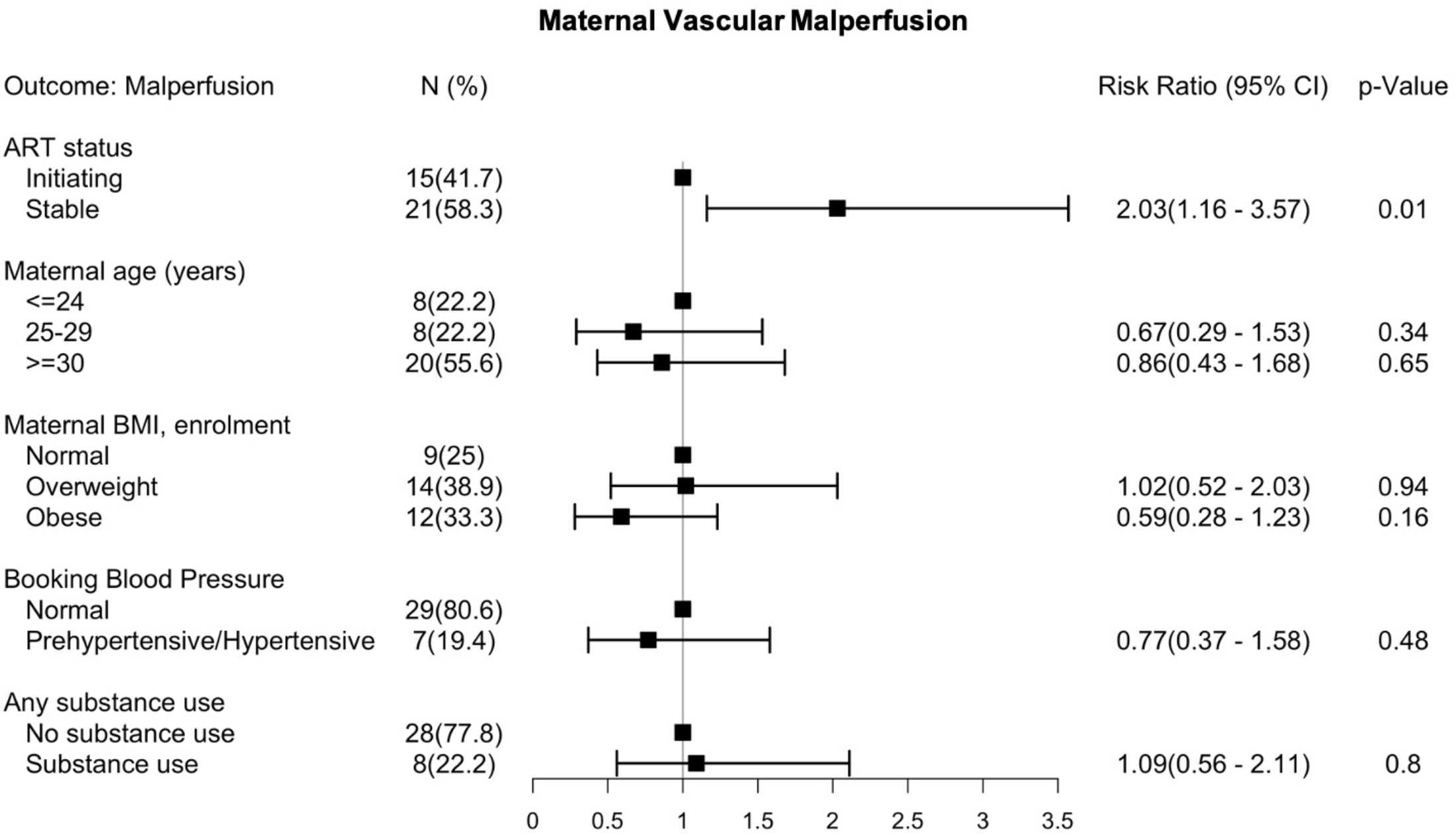
Forest plot depicting the relative risk with the 95% confidence intervals of HIV infected women presenting with placental maternal vascular malperfusion.

**Figure 2D:**
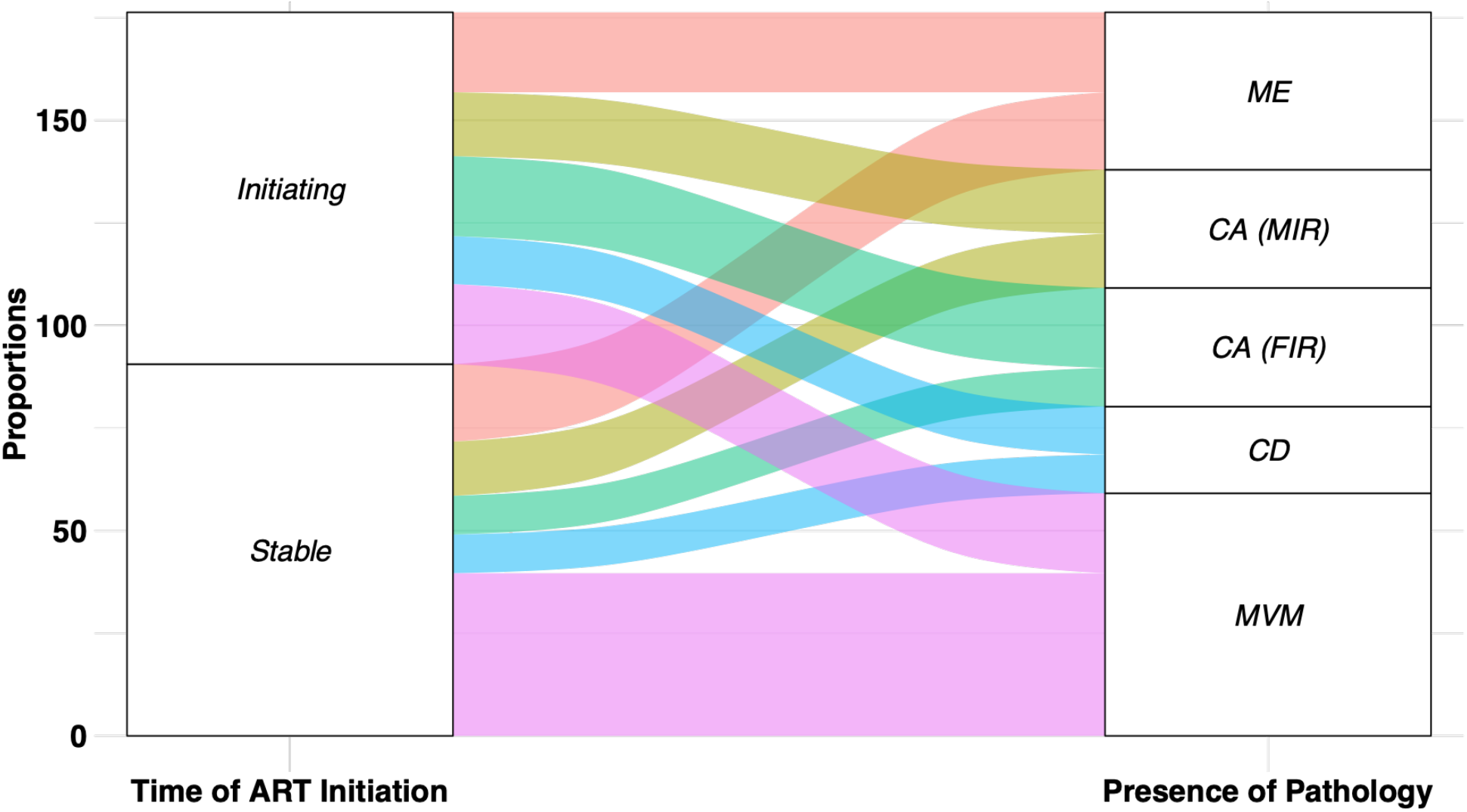
Alluvial plot depicting the impact of ART timing on the placental pathology among women initiating treatment before pregnancy (*Initiating*) or during pregnancy (*stable*). The plot depicts frequencies from a subset of women with abnormal placenta pathology only. These include CA (Chorioamnionitis depicting FIR Fetal Inflammatory Response; MIR, Maternal Inflammatory Response), CD (Chronic Deciduitis), MVM (Maternal Vascular Malperfusion) and Meconium (Meconium Exposure) and the placenta size i.e. <10^th^ percentile for gestational age (small) and > 10^th^ percentile for gestational age (Normal) among women initiating treatment before pregnancy (*Initiating*) or during pregnancy (*stable*).

## Discussion

We examined the association between timing of ART initiation in HIV-positive women and placental pathology, and with subsequent birth outcomes. Placentas from women who had initiated ART pre-conception, but not those initiating during pregnancy, showed significantly increased maternal vascular malperfusion (MVM). There was no evidence for overt maternal or fetal inflammation in either ART group, contrary to our initial supposition. As MVM was significantly associated with PTD and LBW, our findings suggest that associations between ART and adverse birth outcomes may be placenta-mediated, especially so for women initiating ART before pregnancy, where there is antiretroviral drug exposure during placental implantation and development.

Previous studies of HIV-positive women initiating ART before or during pregnancy have focused on quantifying risk of PTD^[1,4,7,38–41]^, SGA^[4,7,41,42]^, LBW^[7,38,39]^, stillbirth^[43]^ and/or neonatal death^[42]^; virologic failure^[44]^; MTCT^[45]^ maternal plasma angiogenic factors^[46]^, and maternal virologic and immunologic responses^[47]^. This is the first study to investigate placenta pathology in association with timing of ART initiation. In our study, women who initiated ART before pregnancy were significantly older, which could have accounted for the observed MVM. However, when we controlled for maternal age, MVM remained significantly associated with pre-conception ART initiation. Thus, ART initiation pre-conception is a substantial risk factor for MVM.

MVM represents a pattern of placental injury which affects the maternal vasculature and circulation and often associates with significantly increased risk of adverse effects to the fetus^[24,29]^. We found that the high incidence of MVM in our study was consistent with findings from a previous HIV-positive pregnancy cohort^[48]^, albeit MVM in our study associated with PTD and LBW. We posit that prolonged ART, initiated pre-conception, is likely resulting in perturbed placental development and poor perfusion and function giving rise to adverse birth outcomes. Interestingly, there was a non-significantly higher proportion of macrosomia births in the women initiating ART during pregnancy, perhaps reflecting the higher proportions of mothers with a high BMI in this group. Although macrosmia has been associated with shoulder dystocia, neonatal and maternal trauma,^[49]^ hypogycaemia,^[50]^ this was not observed in our larger PIMS cohort when we stratified the groups by time of ART initiation (before versus during pregnancy).^[51]^

Although HIV-positive individuals on ART have been shown to have a high prevalence of hypertension^[52]^, women initiating ART prior to pregnancy in our study were less likely to be pre- or hypertensive than those initiating during pregnancy. Additionally, we did not find any associations between MVM and hypertensive disorders in our cohort, although hypertensive disorders of pregnancy, including preeclampsia and chronic hypertension, are the classical maternal clinical features associated with an increased risk of MVM^[53–55]^. Our data suggests that placental research should form an integral part when exploring interventions aimed at mitigating adverse birth outcome associated with ART use in pregnancy.

The impact of ART exposure on the placenta is under-explored and we speculate that exposure of ART before or during early pregnancy results in systemic maternal inflammation that may disrupt vasculogenesis and angiogenesis^[9]^. In our cohort, the underlying mechanism leading to this disruption may be dysregulation of pro-angiogenic mediators including placental growth factor (PIGF), soluble endoglin (sEng) and angiopoietin-1 (Ang-1). Although data is scarce, previous findings demonstrate that maternal HIV infection and ART exposure is associated with an altered pro-angiogenic state; including lower PlGF, higher sEng, decreased Ang-2 and increased Ang-1^[10,56–58]^. Based on previous evidence linking ART with dysregulated angiogenesis and compromised placental vascular development in early pregnancy, we postulate that the higher incidence of MVM in women who initiate ART before pregnancy was likely due to ART-associated shifts towards an anti-angiogenic state ^[59]^. This may account for the increased MVM in women intiating ART pre-conception; suggestive that ART exposure could have detrimental effects on placental development characterized by altered uterine and intervillous blood flow dynamics.

Of potential importance for the interpretation of our findings was that over three quarters of the women were either overweight or obese at first antenatal visit; increased BMI at that time was associated with a decreased likelihood of a small placenta at delivery. We did not have reliable assessments of BMI pre-pregnancy, but in other studies^[53,54]^ elevated pre-pregnancy BMI has been associated with large for gestational age placentas and altered placental metabolic function. We recently showed from our larger PIMS cohort, maternal obesity can increase the risk of having high birthweight and large for gestational age infants and that incremental gestational weight gain increased the risk of having a spontaneous preterm delivery.^[55]^ Beyond the effect of high BMI during pregnancy on the placenta, it is also associated with childhood obesity and cardiovascular diseases in later life.^[56,57]^ Although our study was limited in this regard, interventions targeting the risks associated with the high rate of obesity among women of child-bearing age or during gestation should not be ignored

The main strength of our study is the cohort design allowing us to compare two groups of pregnant women receiving the same NNRTI-based ART regimen and enrolled early in gestation, with data collected and samples investigated according to a strict protocol. Additional strength lies in the ultrasound-based gestational age assessment, leading to accurate PTD determination. Limitations include the non-random nature of the selection of the nested cohort, giving rise to potential differences in maternal characteristics between ART groups or placentas collected. However, we doubt this would affect our findings as: a) there was no significant difference in maternal characteristics when we compared the placenta sub-group with the overall cohort and b) the rates of stillbirths and miscarriages were less than 5% in the larger PIMS cohort, with no differences in pregnancy loss when groups were stratified by time of ART initiation. All staff in the three clinics where women delivered were trained in placenta collection (with no exclusion criteria), and placentas were transported to the laboratory for processing immediately.

In conclusion, we show women receiving ART pre-conception were at increased risk of MVM, suggestive of placental dysfunction. In turn, the association between MVM with PTD and LBW suggests that a placenta-mediated mechanism links the putative association between ART and adverse birth outcomes. Our results further intimate that monitoring placental function in women who initiated ART prior to conception is warranted and that adjunctive therapies that target placental insufficiency in these women can alleviate the burden of adverse birth outcomes.

## Data Availability

The data collected in this study will be available to external investigators interested in collaboration upon submission and approval of a data analysis plan. Requests for data available within PIMS or to submit a request for additional data collection should be submitted to and will be reviewed by the study steering committee

## ACKNOWLEDGEMENTS

We thank all the study participants in this study; and all the members of the PIMS study. We also wish to thank Sue Ford, Colin Newel for data management and Michael Zulu for specimen processing.

## Author contributions

Clive M Gray, Marie-Louise Newell, Thokozile R Malaba, Landon Myer: Conceptualization and design of the study

Thokozile R Malaba, Hlengiwe P Madlala: study conduct, data collection and specimen collection

Nadia M Ikumi: Laboratory and sample preparation

Komala Pillay, Marta Cohen: Histopathology scoring and interpretation

Nadia M Ikumi, Thokozile R Malaba: Statistical analysis

Nadia M Ikumi, Thokozile R Malaba, Clive Gray, Hlengiwe P Madlala, Marie-Louise Newell, Mushi Matjila, Dilly Anumba, Landon Myer: Writing the manuscript, review and editing

---

**Supplementary Table 1:** Substance use among study participants.

|  | **Total**<br>**N=130** | **ART Initiation before**<br>**pregnancy**<br>**N=53** | **ART Initiation**<br>**during pregnancy**<br>**N=77** | **p-Value** |
| --- | --- | --- | --- | --- |
| Substance Use† |  |  |  |  |
| Alcohol |  |  |  | 0.2 |
| Yes | 22 (16.9) | 6 (11.3) | 16 (20.8) |  |
| No | 105 (80.8) | 45 (84.9) | 60 (77.9) |  |
| Missing | 3 (2.3) | 2 (3.8) | 1 (1.3) |  |
| Smoking |  |  |  | 0.1 |
| Yes | 7 (5.4) | 5 (9.4) | 2 (2.6) |  |
| No | 121 (93.1) | 46 (86.8) | 75 (97.4) |  |
| Missing | 2 (1.5) | 2 (3.8) | 0 (0) |  |
| Drugs |  |  |  | 0.5 |
| Yes | 2 (1.5) | 0 (0) | 2 (2.6) |  |
| No | 126 (96.9) | 51 (96.2) | 75 (97.4) |  |
| Missing | 2 (1.5) | 2 (3.8) | 0 (0) |  |

†Substance use is self-reported over the last 30 days
<sup>1</sup>Differences between the two groups were assessed using a two-sample t-test or Wilcoxon rank-sum test depending on the distribution for numerical data and the $\chi^2$ test or Fisher's exact test for categorical data.

**Supplementary Table 2:**
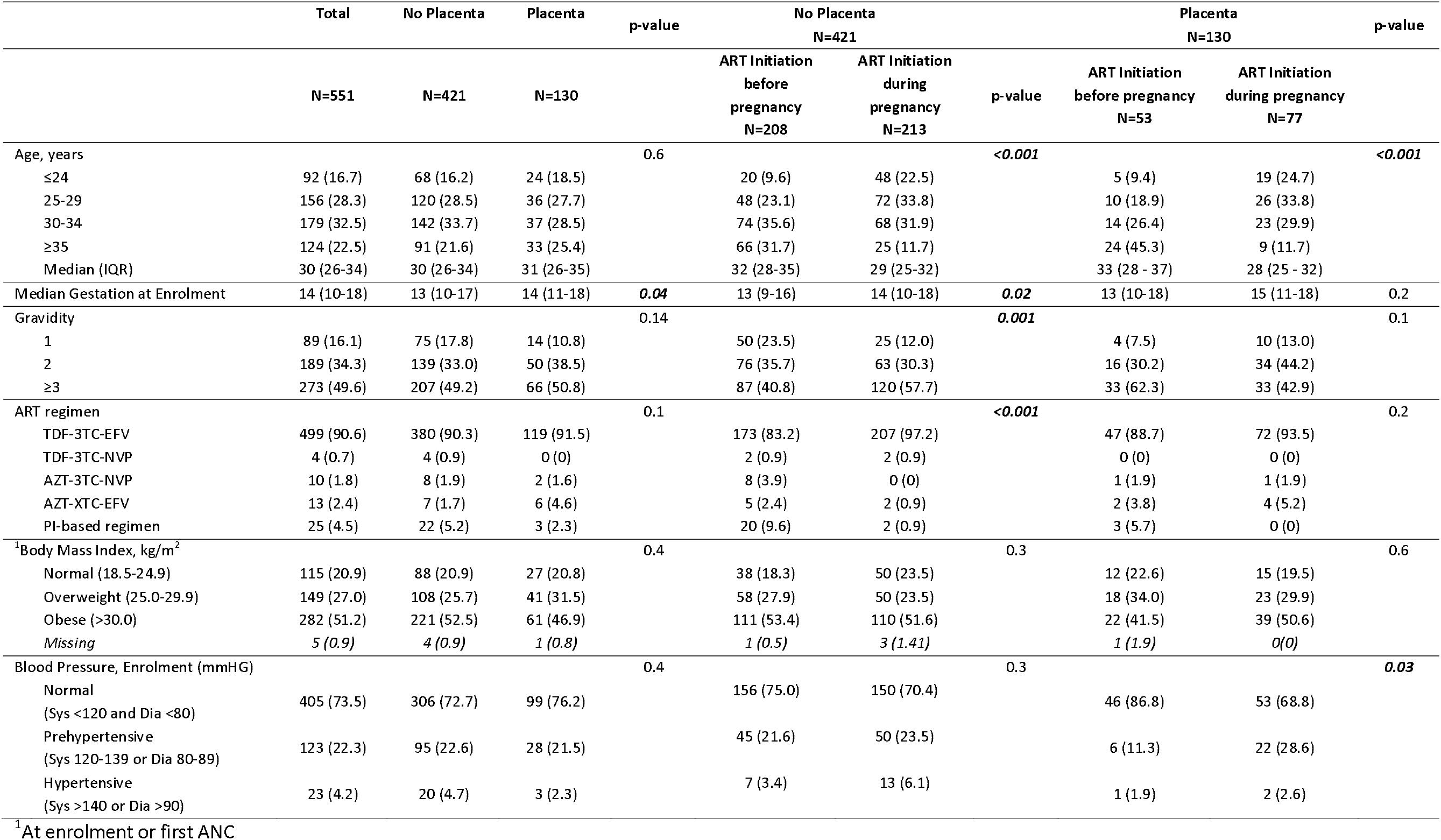
Maternal and obstetric characteristics of pregnant women at enrolment: comparing no placenta vs placenta collection.

